# From Sleep Patterns to Heart Rhythms: Predicting Atrial Fibrillation from Overnight Polysomnograms

**DOI:** 10.1101/2024.06.04.24308444

**Authors:** Zuzana Koscova, Ali Bahrami Rad, Samaneh Nasiri, Matthew A. Reyna, Reza Sameni, Lynn M. Trotti, Haoqi Sun, Niels Turley, Katie L. Stone, Robert J. Thomas, Emmanuel Mignot, Brandon Westover, Gari D. Clifford

## Abstract

**Background:** Atrial fibrillation (AF) is often asymptomatic and thus under-observed. Given the high risks of stroke and heart failure among patients with AF, early prediction and effective management are crucial. Importantly, obstructive sleep apnea is highly prevalent among AF patients (60–90%); therefore, electrocardiogram (ECG) analysis from polysomnography (PSG), a standard diagnostic tool for subjects with suspected sleep apnea, presents a unique opportunity for the early prediction of AF. Our goal is to identify individuals at a high risk of developing AF in the future from a single-lead ECG recorded during standard PSGs.

**Methods:** We analyzed 18,782 single-lead ECG recordings from 13,609 subjects at Massachusetts General Hospital, identifying AF presence using ICD-9/10 codes in medical records. Our dataset comprises 15,913 recordings without a medical record for AF and 2,056 recordings from patients who were first diagnosed with AF between 1 day to 15 years after the PSG recording. The PSG data were partitioned into training, validation, and test cohorts. In the first phase, a signal quality index (SQI) was calculated in 30-second windows and those with SQI*<*0.95 were removed. From each remaining window, 150 hand-crafted features were extracted from time, frequency, time-frequency domains, and phase-space reconstructions of the ECG. A compilation of 12 statistical features summarized these window-specific features per recording, resulting in 1,800 features. We then updated a pre-trained deep neural network and data from the PhysioNet Challenge 2021 using transfer-learning to discriminate between recordings with and without AF using the same Challenge data. The model was applied to the PSG ECGs in 16-second windows to generate the probability of AF for each window. From the resultant probability sequence, 13 statistical features were extracted. Subsequently, we trained a shallow neural network to predict future AF using the extracted ECG and probability features.

**Results:** On the test set, our model demonstrated a sensitivity of 0.67, specificity of 0.81, and precision of 0.3 for predicting AF. Further, survival analysis for AF outcomes, using the log-rank test, revealed a hazard ratio of 8.36 (p-value of 1.93 × 10^−52^).

**Conclusions:** Our proposed ECG analysis method, utilizing overnight PSG data, shows promise in AF prediction despite a modest precision indicating the presence of false positive cases. This approach could potentially enable low-cost screening and proactive treatment for high-risk patients. Ongoing refinement, such as integrating additional physiological parameters could significantly reduce false positives, enhancing its clinical utility and accuracy.

## I. Introduction

Atrial fibrillation (AF) is one of the most prevalent cardiac arrhythmias in the United States. It is often asymptomatic and therefore under-diagnosed in the general population. It is associated with a higher risk of stroke (5-fold) together with a higher risk of heart failure (3-fold) [1]. Moreover, there is strong evidence of a relationship between AF and obstructive sleep apnea (OSA). Cardiovascular disease and OSA are closely linked with respect to their pathophysiology and epidemiology [2]. The atrial arrhythmogenic mechanism is based on imbalances of the autonomic nervous system, chronic intermittent hypoxia, inflammation, and swings in thoracic pressure during different phases of the acute apnoeic episodes [3]. OSA is acknowledged as a contributing risk factor for the onset and advancement of AF. Additionally, OSA diminishes the efficiency of antiarrhythmic medications, electric cardioversion (EC), and catheter ablation (CA) procedures in managing AF [4]. The gold standard for diagnosis of OSA in older individuals and those with cardiac and pulmonary comorbidities is laboratory polysomnography (PSG). PSG encompasses the simultaneous recording and analysis of various physiological parameters during sleep, including electroencephalography (EEG), electrooculography (EOG), electromyography (EMG), monitoring respiratory effort, airflow dynamics, blood oxygen saturation levels as well as electrocardiography (ECG). Those undergoing PSG are at higher risk for AF than those who have home sleep apnea tests (which often do not record ECG). This gives a unique opportunity to predict future AF from the ECG channel given the higher risk of AF for patients with suspected sleep apnea.

## II. Dataset

In this study, we utilized data from the Massachusetts General Hospital collected for the Human Sleep Project [5]. Our dataset comprises 18,782 PSG recordings from 13,609 patients. All signals are measured in microvolts with a sampling frequency of 200 Hz. For the prediction of future AF, a single ECG channel recorded below the right clavicle near the sternum and over the left lateral chest wall was utilized. The mean length of the ECG recording is 7.62 ± 1.34 hours.

We retrieved the AF vs. AF-free labels together with the date of AF being diagnosed from electronic health records (EHR) based on the ICD9 (427.31, 427.32) and ICD10 (I48.0, I48.1, I48.2, I48.91) codes [6], [7]. For identifying incident AF from EHR, we used the following definition: Two ICD 9/10 codes (out or inpatient) separated by more than seven days but within one year [7]. Table I summarizes the number of recordings for three different categories: AF-free, AF-past pointing to the patients diagnosed with AF prior to their PSG date, and AF-future representing patients diagnosed with AF after the PSG recording.

**TABLE I.** Subjects and recordings distribution by the presence of AF.

| Data | Subjects | Recordings |
| --- | --- | --- |
| AF-free | 11,898 | 15,913 |
| AF-past | 327 | 413 |
| AF-future | 1,704 | 2,456 |

For the prediction of future AF, we will continue to use recordings that are either from the category AF-free or AF-future, including only patients who were first diagnosed with AF from 1 day to 15 years after the day PSG was taken. The progression of AF is captured by a swimmer plot (Figure 1) representing the time elapsed from the PSG date to the diagnosis of AF in the EHR.

**Fig. 1.**
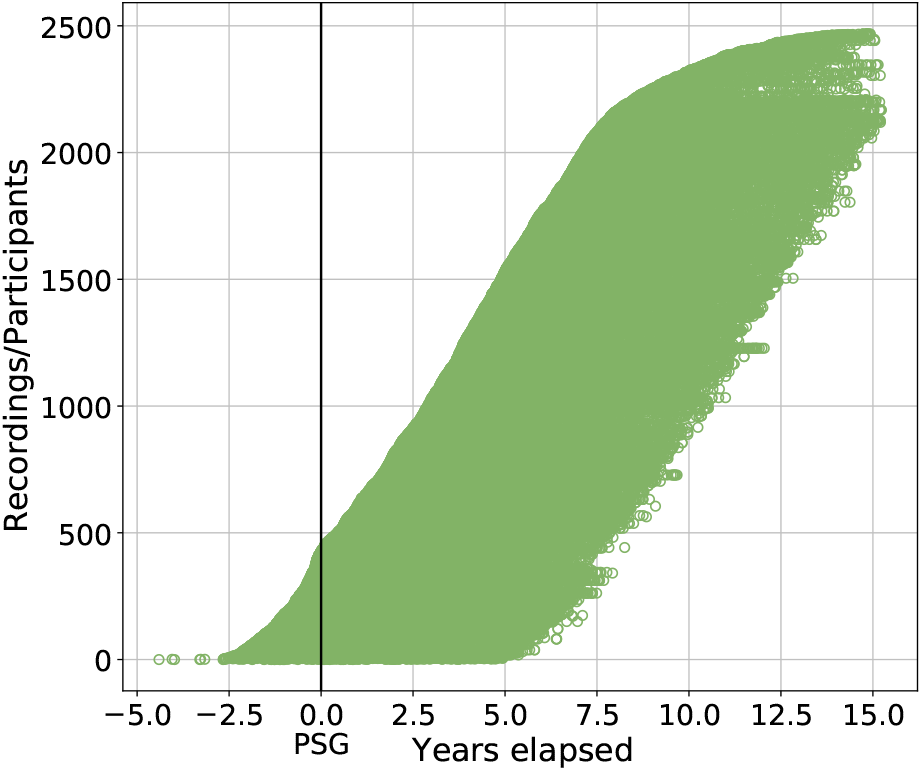
Swimmer plot illustrating the progression of AF events for individual participants. The vertical line marks the time the PSG was conducted, while each horizontal circle represents the duration from the PSG date to the diagnosis of AF in EHR.

## III. Methods

We employed two techniques to predict future AF: an algorithm for feature extraction and a deep learning method to determine the probability of AF, which were then combined into a single prediction algorithm.

### A. Feature extraction algorithm

The algorithm for feature extraction is based on the winning solution [8] from the PhysioNet Challenge 2017 focused on the classification of AF from short single lead ECG recordings [9]. The processing pipeline consists of upsampling the ECG to 300 Hz, splitting the ECG into 30-second windows to align with sleep stages, and consequently calculating the signal quality index (SQI) by comparing the results of two different peak detection algorithms. The 30 s window is discarded from further analysis in case SQI is lower than 0.95. Segments with satisfying SQI (higher than 0.95) are denoised and the baseline wander is removed based on the sparse derivative decomposition algorithm [10]. Subsequently, a set of 150 hand-crafted ECG features is extracted containing time domain and morphological features, frequency domain features, time-frequency domain features, and nonlinear (phase space) features [8]. This results in an array of size N x 150, where N represents the number of 30-second segments of the recording. Subsequently, 12 statistical features (mean, median, standard deviation, skewness, kurtosis, maximum, minimum, mean gradient, median gradient, maximum gradient, minimum gradient and standard deviation of the gradient) are extracted for each one of ECG hand-crafted features across N-30s windows resulting in 1,800 (12 statistical features x 150 ECG hand-crafted features) ECG statistical features.

### B. Deep learning approach

The second approach uses the deep learning solution [11] from the PhysioNet Challenge 2021 [12] focused on arrhythmia classification with emphasis on differences in performance for varying numbers of input ECG channels. The model architecture is designed as a custom Residual neural network with an attention layer, pre-trained on 86,000 recordings from the public PhysioNet Challenge 2021 dataset for 26 arrhythmias classification. We then updated a pre-trained deep neural network using transfer learning to discriminate between recordings with and without AF using the same Challenge data by changing the last fully connected layer.

During the preprocessing stage, the PSG’s ECG recordings are upsampled to 500 Hz, bandpass filtered from 1–47 Hz, divided into 16-second windows (8,192 samples), and each 16-second window is standardized using z-score. The pre-trained model was then applied to the 16-second windows to generate the probability of AF. The result is the *M* × 1 probability vector, where *M* is the number of 16-second windows across the recording. Subsequently, we extracted the same 12 statistical features out of the probability vector and added a 13th feature; the number of segments with a probability higher than threshold (0.45). The threshold was estimated empirically on the PhysioNet Challenge 2021 training dataset.

### C. Final prediction model

In the final phase, we combined 1,800 features extracted from our feature extraction algorithm with an additional 13 features derived from the posterior probability of the deep learning model. The PSG data were partitioned into training, validation, and test sets (Table II) ensuring that individual patients remained exclusive to each cohort. Stratification based on the target label was applied across all cohorts.

**TABLE II.** Recordings partition into the training, validation, and test sets.

|  | Training | Validation | Test |
| --- | --- | --- | --- |
| AF-free | 12,903 | 1,400 | 1,610 |
| AF-future | 1,668 | 193 | 195 |

Subsequently, we trained a random forest classifier consisting of 200 trees and computed feature importance by assessing the mean and standard deviation of impurity decrease accumulation within each tree. Through grid search, we identified the top 1,000 most significant features. Following this, we constructed a neural network (NN) architecture comprising 5 blocks, each containing identical layers with varying hyperparameters: a fully connected layer with ReLU activation function, batch normalization layer, and dropout layer. The output dimensions of these layers are 128, 128, 64, 32, and 2, respectively, with a dropout probability of 0.2 for all layers except the last, where it is set to 0.5. A softmax activation function is appended after the final block to yield the probability of AF-free versus AF-future. Throughout the training, we employed the Adam optimizer with a learning rate of 0.001 and incorporated a learning rate scheduler, adjusting the rate dynamically with a step size of 7 epochs and a gamma value of 0.1 over a training span of 32 epochs to optimize convergence. After training the model, we determined the optimal threshold for class dichotomization as the operating point of the receiver-operating characteristic (ROC) curve, set at 0.59. Subsequently, we evaluated the model’s performance on the test cohort.

### D. Analysis of performance for different time frames

After training the model, we assessed the model’s performance across various time windows ranging from one to fifteen years for the test set. To ensure accurate evaluation, we employed point censoring techniques. Point censoring arises when, despite ongoing monitoring, patients are lost to follow-up or the event of interest does not occur within the study’s time frame. More specifically, for patients free of AF, we censored those whose observation period was shorter than a defined time frame. Conversely, for patients categorized as AF-future, we censored individuals diagnosed with AF after the specified time frame. After applying censoring, we assessed the model’s sensitivity, specificity, and positive predictive value (PPV) across follow-up time frames ranging from 1 to 15 years, with a step size of 1 year.

### E. Survival analysis

The model’s ability to stratify patients into high-risk and low-risk groups for future AF was assessed using survival analysis on the test set. To measure the relative risk of AF occurrence between groups over time we computed the hazard ratio (HR) using the log-rank test. We conducted survival analysis by selecting the initial recording and its corresponding prediction from each subject, as some subjects contributed multiple recordings.

### F. Sleep apnea, stroke and AF association

To test the association between sleep apnea and future AF we conducted a chi-squared test on the test cohort. Sleep apnea labels were extracted from PSG recording annotation files, and the apnea-hypopnea index (AHI) was calculated. Patients with an AHI higher than 5 were categorized as having sleep apnea [13].

Subsequently, we calculated the chi-squared statistic to assess the difference between the observed and expected frequencies, assuming independence between sleep apnea and atrial fibrillation.

We also conducted a chi-squared test to examine the association between future stroke and predicted future AF. Stroke labels were obtained from EHR based on ICD9 (430, 431, 433, 434.9, 434, 436) and ICD10 codes (I60, I61, I63, I64, I63.9) [14].

## IV. Results

The overall results of future AF prediction for both the validation and test datasets are provided in Table III. While the performance regarding AUROC is relatively impressive at 0.82 on both validation and test data, the PPV and AUPRC show modest performance.

**TABLE III.** Results for validation and test sets for future af prediction.

| Metric | Validation | Test |
| --- | --- | --- |
| AUROC | 0.82 | 0.82 |
| AUPRC | 0.46 | 0.42 |
| PPV | 0.34 | 0.30 |
| Sensitivity | 0.70 | 0.67 |
| F1 | 0.46 | 0.42 |
| Specificity | 0.81 | 0.81 |

The results for different time windows after point censoring are captured in Figure 2 with the corresponding counts of recordings for each time frame captured in Table V.

**Fig. 2.**
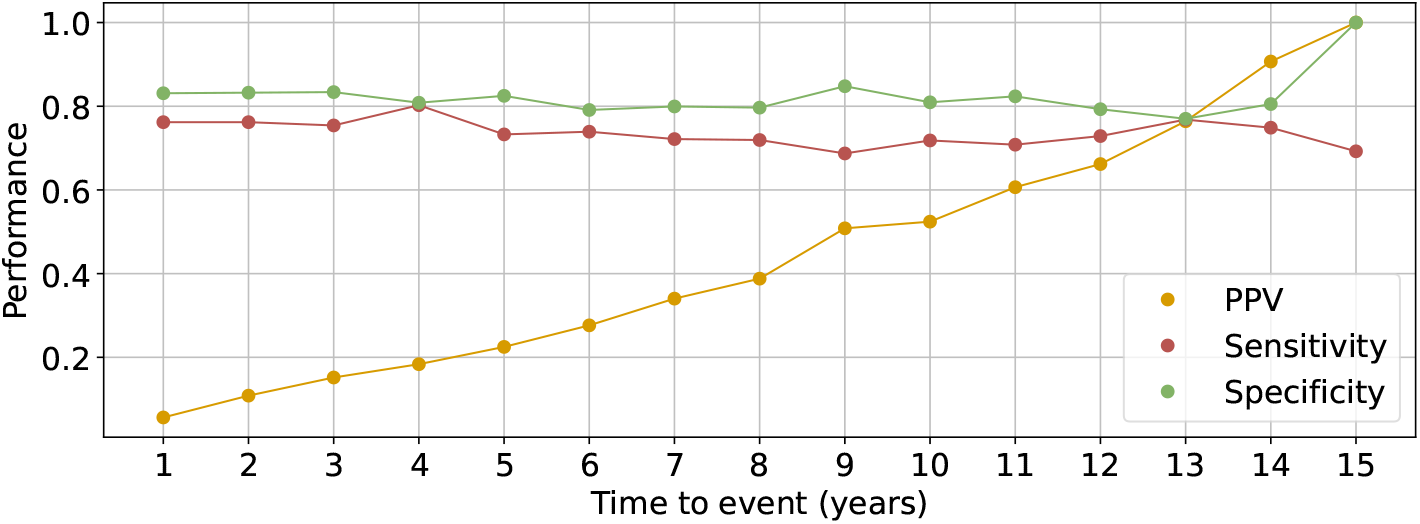
Illustration of model performance for future AF prediction for different time windows ranging from 1-15 years. The illustration encompasses sensitivity, PPV, and specificity metrics. Additionally, Table V indicates the follow-up time in years in the first row, while the second and third rows represent the corresponding counts of recordings for AF-free and AF-future categories after applying point censoring.

### A. Survival analysis

In the context of survival analysis, we obtained an HR of 8.36 (95% CI: 5.60 to 12.48) with *p*-value of 1.93 × 10^−52^. The Kaplan-Meier plot illustrating these findings is depicted in Figure 3.

**Fig. 3.**
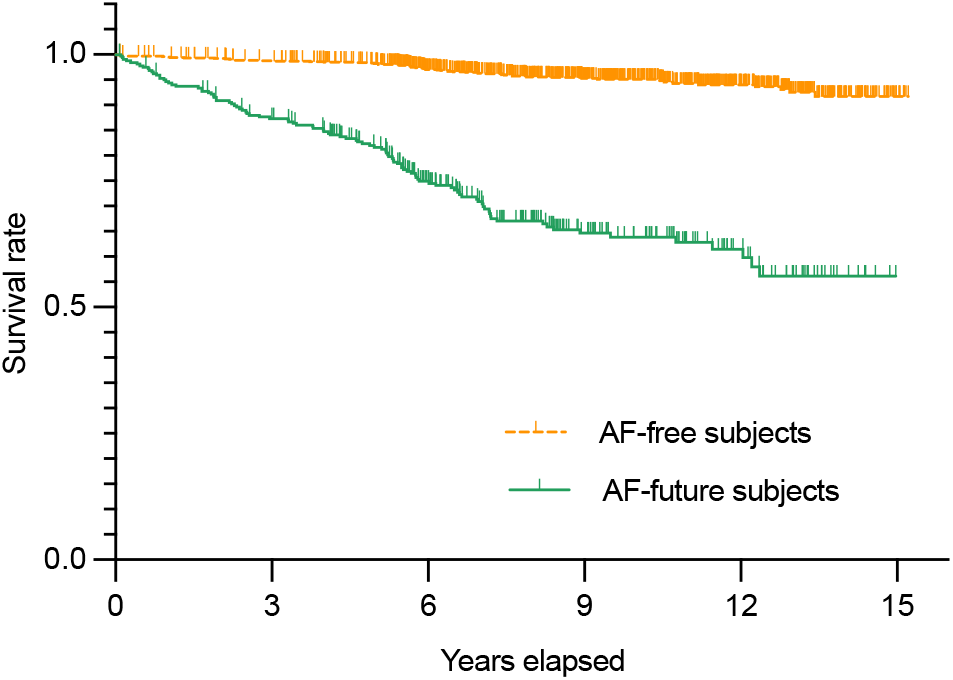
The Kaplan-Meier curve illustrates the probability of survival with outcome AF for two distinct groups. One group comprises subjects classified by the model as AF-free represented by the orange curve, while the other group consists of individuals deemed to be at high risk of developing AF in the future depicted by the green curve. The log-rank test revealed a hazard ratio of 8.36 (95% CI: 5.60 to 12.48), indicating a significant difference between the groups (p *<*0.0001).

### B. Sleep apnea, stroke and AF association

The contingency tables summarizing the observed and aexpected frequencies for the presence or absence of sleep apnea and AF are presented in Table IV.

**TABLE IV.** Contingency table of observed and expected frequencies of records for apnea vs predicted af.

| Presence of Apnea | Values | AF-future | AF-free |
| --- | --- | --- | --- |
| Apnea | Observed | 304 | 696 |
| Apnea | Expected | 240 | 760 |
| Apnea-free | Observed | 113 | 623 |
| Apnea-free | Expected | 177 | 560 |

**TABLE V.** Number of recordings after point censoring for different time frames.

| Time (years) | 1 | 2 | 3 | 4 | 5 | 6 | 7 | 8 | 9 | 10 | 11 | 12 | 13 | 14 | 15 |
| --- | --- | --- | --- | --- | --- | --- | --- | --- | --- | --- | --- | --- | --- | --- | --- |
| AF-free | 1,586 | 1,569 | 1,546 | 1,508 | 1,456 | 1,278 | 1,102 | 954 | 782 | 619 | 482 | 338 | 200 | 77 | 10 |
| AF-future | 21 | 42 | 61 | 81 | 101 | 138 | 158 | 171 | 179 | 181 | 185 | 188 | 194 | 195 | 195 |

Our chi-squared test yielded a statistic of 51.77 and a *p*-value of 6.23 × 10^−13^, indicating a significant association between sleep apnea and future atrial fibrillation.

The observed and expected frequencies for the presence or absence of future stroke and AF are displayed in Table VI. The chi-squared test statistic yielded 20.9, with a *p*-value of 4.85 × 10^−6^, indicating a significant association between future stroke and predicted future AF.

**TABLE VI.** Contingency table of observed and expected frequencies for future stroke vs predicted af.

| Future stroke | Values | AF-future | AF-free |
| --- | --- | --- | --- |
| Stroke | Observed | 72 | 120 |
| Stroke | Expected | 46 | 146 |
| Stroke-free | Observed | 360 | 1,253 |
| Stroke-free | Expected | 386 | 1,227 |

## V. Discussion

The results of our method for predicting future AF on the test cohort demonstrate promising performance across various evaluation criteria as illustrated in Table III. The specificity of 0.81 indicates a satisfactory ability of the model to identify patients as AF-free in the future. Moreover, a sensitivity of 0.67 signifies the model’s acceptable capability in detecting patients at high risk of developing AF. While the PPV stands at 0.3, suggesting room for improvement due to a notable number of false positive cases, it is essential to contextualize this metric. PPV is a prevalence prevalence-dependent metric, and exceeds the prevalence of 15% for future AF in our dataset.

Given our time-to-event data, we aimed to assess our algorithm’s performance in predicting future AF across various time frames spanning 1 to 15 years. Identifying patients at risk of developing AF within the initial years following the PSG is crucial for timely intervention and treatment strategies. As depicted in Figure 2, our observations reveal a slight decrease in sensitivity over the 15-year prediction period. Conversely, specificity remains relatively stable throughout the duration, showing no significant fluctuations. Notably, we observe an improvement in PPV with an increasing time window reflecting the influence of class prevalence.

The survival analysis for AF outcome, visually represented by the Kaplan-Meier plot in Figure 3, unveiled a significantly higher survival rate for subjects predicted by our model as low risk for developing AF. The hazard ratio of 8.36 indicates that the rate of AF occurring in the group predicted as high risk of having AF is more than 8 times the rate of the AF-free group. This study also aimed to investigate the relationship between current sleep apnea and the predicted future AF. Upon comparing the observed frequencies in the dataset with the expected frequencies for sleep apnea and AF using the chi-squared test (Table IV), we determined a significant association (*p*-value *<*0.0001). These findings underscore the critical importance of AF screening for patients undergoing a PSG, given the shared pathophysiology between sleep apnea and AF.

Furthermore, the significance of screening patients for future AF is highlighted by the findings of the chi-squared test examining the association between predicted AF and future stroke diagnostic labels (Table VI). Moreover, our model predicted AF for 37.5% of all stroke incidences, suggesting that preventative screening could potentially reduce the incidence of future strokes.

While the results are tantalizing for mass AF risk screening (over 2 million PSGs are performed yearly in the USA), it is important to note some limitations of our study. First, since the volume of ECG was vast (over 140,000 hours of data) it was unfeasible to overread the ECGs to identify if the algorithm was detecting or predicting the presence of AF. In future work we aim to randomly sample the ECG with high probabilities of AF, to identify undiagnosed episodes. Nevertheless, it might be problematic to definitely classify a patient as having AF just from some random samples. Another option for identifying subjects with AF already present might be running multiple existing AF classification algorithms, validated on the external datasets. By focusing on segments classified as AF with high algorithmic consensus, we can categorize these recordings into a group suggestive of present but undiagnosed AF.

Regardless, this may just be semantics, since no longitudinal study of AF has been performed which identifies how AF evolves in an unselected population. It may be that patients that go on to be diagnosed as suffering from AF, were already exhibiting some AF episodes ’silently’. In that sense, there would be no real ’prediction’ of AF, but just early identification. Of course, one may argue this for almost all diseases, since the definitive diagnosis is often just defined as passing a certain threshold.

For future AF, it is possible that patients who were identified as having a high risk of future AF did already manifest AF at some point, but it was either in another health system or went unnoticed. This issue would only serve to reduce our model’s performance.

Finally, we note that cardiac dynamics during sleep are complex, and not well understood. Sleep influences the cardiovascular, endocrine, and thermoregulatory systems, and the circadian patterns of blood pressure, and sympathetic and parasympathetic activation change throughout the night. Thus, sleep stage (NREM/REM, deep/light NREM) or state (stable/unstable) specific analysis may reveal more mechanisms and predictive power for our model.

## VI. Conclusion

This study introduced a novel method for predicting future AF using single-lead ECG recordings from standard PSG data. Our approach leverages a large dataset of ECG recordings, demonstrating the capacity to pinpoint individuals at elevated risk of AF years before clinical manifestation. The model exhibited acceptable sensitivity and specificity within the test cohort, with a significant hazard ratio identified through survival analysis. However, the PPV indicates room for improvement in reducing false positives.

These results suggest the potential of our method as a non-invasive screening tool that could be integrated into existing diagnostic protocols for sleep apnea, enhancing early detection and management of AF. Ongoing refinement and rigorous validation are essential to improve accuracy and clinical utility. With further development, this approach could significantly impact AF management, improving patient outcomes and reducing healthcare costs associated with its complications.

## Data Availability

All data produced in the present study are available upon reasonable request to the owners of the Human Sleep Project https://bdsp.io/content/hsp/1.0/

## VII. Acknowledgements

This study was funded by the National Heart, Lung, and Blood Institute (NHLBI) under NIH grant R01 HL161253.

## Notes

### Competing Interest Statement

The authors have declared no competing interest.

### Author Declarations

https://bdsp.io/content/hsp/1.0/

